# Neck pain service utilization and costs: association with number of visits of chiropractic manipulation, active care, manual therapy, or acupuncture. A retrospective cohort study

**DOI:** 10.1101/2023.01.12.23284483

**Authors:** David Elton, Meng Zhang

## Abstract

**Background:** Neck pain (NP) clinical practice guidelines (CPG) generally emphasize natural history, self-care, and non-pharmaceutical therapies. For non-pharmaceutical therapies provided for NP, like chiropractic manipulative treatment (CMT), active care (AC), manual therapy (MT), or acupuncture, little is known about the dose/response relationship with use of other services and total cost. The purpose of this retrospective cohort study of individuals with NP was to examine the dose response association between the number of visits of CMT, AC, MT, or acupuncture, the exposure to pharmaceutical, imaging, and interventional services, and total episode cost.

**Methods:** Episode of care was used to analyze a national sample of individuals 18 years and older with a single episode of non-surgical NP occurring in 2017-2019 and initially contacting a chiropractor (DC), physical therapist (PT), or licensed acupuncturist (LAc). The number of visits of CMT, AC, MT, or acupuncture were the primary independent variables. Rate and timing of use of 13 types of health care services and total episode cost were the primary dependent measures.

**Results:** A total of 91,805 continuously insured individuals initially contacted a DC, PT, or LAc for a single episode of non-surgical NP. These individuals initially contacted 19,387 different DCs, 1,828 PTs and 1,153 LAcs. There were $39,150,944 in total expenditures. The most common number of visits was 1 to 3 for CMT (47.8% of episodes), AC (31.8%), and MT (35.0%), and 4 to 6 for acupuncture (27.5%). Different levels of utilization intensity of CMT, AC, MT, and acupuncture were generally not associated with statistically or clinically meaningful differences in exposure to pharmaceutical, imaging, or interventional services. Total episode cost increased with higher numbers of visits of CMT, AC, MT, and acupuncture with CMT associated with the lowest median total episode code at each level of visit utilization.

**Conclusions:** For individuals with non-surgical NP initially contacting a DC, PT or LAc, 1 to 3 visits of CMT, AC, or MT, and 4 to 6 visits of acupuncture were the most common levels of utilization. A higher number of visits of CMT, AC, MT or acupuncture was associated with significantly higher total cost, without clinically or statistically meaningful differences in exposure to pharmaceutical, imaging, or interventional services. CMT was associated with the lowest total episode cost at each level of utilization. Higher visit counts of CMT, AC, MT, or acupuncture may have been associated with unmeasured clinical benefits and warrants further study.

## Background

Neck pain (NP) is prevalent ^1,2^ and costly.^3^ Among spinal disorders, in the absence of red flags of serious pathology low back pain (LBP) clinical practice guidelines (CPGs) describe a stepped approach in which services are sequenced into first-, second- and third-line services.^4-6^ NP CPGs, while less abundant and more heterogeneous than those for LBP, are similar emphasizing individual self-management, non-pharmacological and non-interventional services as first-line approaches.^7-9^

Variation in service utilization and cost outcomes for NP has been associated with the initial contact health care provider (HCP).^10-14^ Like LBP ^15^, when initially contacted by an Individual with NP, non-prescribing HCPs, like chiropractors (DC), physical therapists (PT), or licensed acupuncturists (LAc) are more likely to have episodes involving non-pharmacological and non-interventional therapies and less use of pharmaceutical, imaging, and interventional services.^14^

Among spinal disorders, dose response analyses of chiropractic manipulative treatment (CMT), active care (AC), and manual therapy (MT) are more common for LBP than for NP.^16-21^ LBP dose response analyses have not found a clear association between a higher dose and beneficial impact on cost, quality adjusted life years, disability and pain free days, and other measures of pain and function.^19-22^ Dose response analyses of CMT, AC and MT for NP are less common ^18,23,24^, with dose response analyses of acupuncture even less common.^25^ In a cohort with insurance coverage, an important consideration for analyzing dose/response is patient willingness to pay (WTP) for the co-payments, deductibles or coinsurance associated with CMT, AC, MT, or acupuncture services.^26^

For individuals with NP initially contacting a DC, PT or LAc the aim of this study was to examine the association between the number of visits of CMT, AC, MT, or acupuncture services, utilization of other health care services, and total cost. The hypothesis was that increasing the number of visits of these services would increase total episode cost and have minimal impact on utilization of pharmaceutical, imaging, and interventional services.

## Methods

### Study design, population, setting and data sources

This was a retrospective cohort study of individuals with a single episode of non-surgical NP during 2017-2019 for which the individual initially contacted a DC, PT or LAc. The study design was identical to a previous LBP study.^27^ An enrollee database included deidentified enrollment records, and administrative claims data for all inpatient and outpatient services, and pharmacy prescriptions for enrollees from a single national commercial insurer. Deidentified HCP demographic information and professional licensure status was included in an HCP database. ZIP code level household adjusted gross income (AGI) was extracted from the Internal Revenue Service ^28^, population race and ethnicity from the US Census Bureau ^29^, socioeconomic status (SES) Area Deprivation Index (ADI) data, from the University of Wisconsin Neighborhood Atlas^®^.^30^

With study data de-identified or a Limited Data Set in compliance with the Health Insurance Portability and Accountability Act and customer requirements, the UnitedHealth Group Office of Human Research Affairs determined that this study was exempt from Institutional Review Board review. The study was conducted and reported based on the Strengthening the Reporting of Observational Studies in Epidemiology (STROBE) guidelines (Supplement – STROBE Checklist).^31^

With numerous potential unknown or unmeasurable confounders, and to avoid creating any causal impressions, the analysis did not adopt standard practice to adjust for typical confounders such as individual age, sex and co-morbidities ^32,33^ using common yet potentially inadequate approaches such as propensity score matching.^34^ Potential confounders of CMT, AC, MT and acupuncture visit intensity include: anticipated potential out of pocket costs and WTP; time availability to participate in multiple visits; DC, PT or LAc options convenient to an individual’s home, workplace or daily travel routes and mode of transportation; individual preference type of HCP including gender or racial concordance; recommendations from family or friends; perceived NP severity; and appointment availability within an individual’s timing expectations meeting these and other criteria.^35^ Actual measures of individual demographic and episodic characteristics, and related associations, were calculated for different levels of visit dose for each service type.

### Cohort selection and unit of analysis

To match the timeframe used in an identical study of LBP ^27^, and to avoid the influence of the COVID-19 epidemic on care patterns in early 2020, the cohort included individuals aged 18 years and older with a single complete episode of NP commencing and ending during the calendar years 2017-2019. All individuals had continuous medical and pharmacy insurance coverage during the entire study period. The study cohort was able to access DC, PT, and LAc HCPs directly without a referral.

The Symmetry^®^ Episode Treatment Groups^®^ (ETG^®^) and Episode Risk Groups^®^ (ERG^®^) version 9.5 methodologies and definitions were used to translate administrative claims data into episodes of care, which have been reported as a valid measurement for comparison of HCPs based on cost of care.^36^ This approach has been shown to be a valid way to organize all administrative claims data associated with a condition.^37^ The risk of misclassification bias associated with using episode of care as the unit of analysis has previously been demonstrated to be low.^14,15,27^

With the cohort having continuous insurance coverage, complete episodes were identified as having at least 91-day pre- and 61-day post-episode clean periods during which no services were provided by any HCP for any NP diagnosis. To minimize the influence of complex NP episodes on study results several exclusions were made including episodes including a surgical procedure or associated with diagnoses of fractures and other spinal trauma, congenital deformities and scoliosis, malignant and non-malignant neoplasms, infection, autoimmune disorders, osteoporosis, or advanced arthritis.

### Variables

Python (*Python Language Reference, Version 3.7.5*., n.d.) was used for data preprocessing, table generation, and initial analyses. A goodness-of-fit measure using D’Agostino’s K-squared test was used to evaluate whether measures were derived from a normally distributed sample. Median and interquartile range (IQR) were used to report non-normally distributed data. Simple descriptive statistics were generated for main study variables.

Of the independent variables of type of HCP initially contacted, DC, PT, or LAc HCP, and the number of visits of CMT, AC, MT, or acupuncture services, the analyses focused on services provided for at least 50% of episodes. This resulted in 4 combinations of HCP and service type: DC-CMT, PT-AC, PT-MT, and LAc-acupuncture (Supplement 1).

The percent of episodes including 13 types of health care services was the primary dependent variable. For the percent calculation any type of HCP an individual saw during the episode of NP could provide a service. To facilitate comparison of NP and LBP^4^, and while NP CPGs do not segment health care services, services were categorized as first-, second-, or third-line using an identical approach as a previous LBP study. ^27^ Total cost of care for all reimbursed services provided by any HCP during an episode, the number of different HCPs seen during an episode, and episode duration measured in days were secondary dependent variables. Total episode cost included costs associated with all services provided for NP during an episode, including those not specifically identified in the 13 categories used in the analyses. Costs for services for which an insurance claim was not submitted were not available.

Bivariate analyses, risk ratios (RR) and 95% confidence intervals (CI) were used to evaluate differences in secondary measures compared to the 1 to 3 visit reference for each HCP service type combination. RR were selected instead of odds ratios (OR) due to being more widely understood in associational analyses and because ORs tend to exaggerate risk in situations where an outcome is relatively common.^38^

## Results

19,387 DCs, 1,828 PTs, and 1,153 LAcs were initially contacted by 91,805 individuals with subsequent NP episodes associated with $39,150,944 in reimbursed health care expenditures. While pre- and post-episode clean periods were similar, differences were observed in the attributes of individuals initially contacting DCs, PTs, and LAcs. LAcs were initially contacted by 70.5% females, and by individuals from ZIP codes with lower ADI (median 23), higher AGI (median 91,222) and lower % non-Hispanic white (NHW) population (median 63.8%). PTs were initially contacted by slightly older individuals with higher ERG® Risk Score. DCs were initially contacted by individuals from ZIP codes with higher ADI (median 43), lower AGI (median 67,793), and higher % NHW population (median 76.8%) (Supplement 2).

47.8% of DC-CMT episodes, 31.8% of PT-AC, and 35% of PT-MT had 1 to 3 visits. For LAc-acupuncture, 4 to 6 visits were most common, at 27.5% of episodes (Figure 1). Within each HCP and service type combination, similar characteristics were found in individual, population, and episode attributes among visit count categories (Supplement 3).

**Figure 1.**
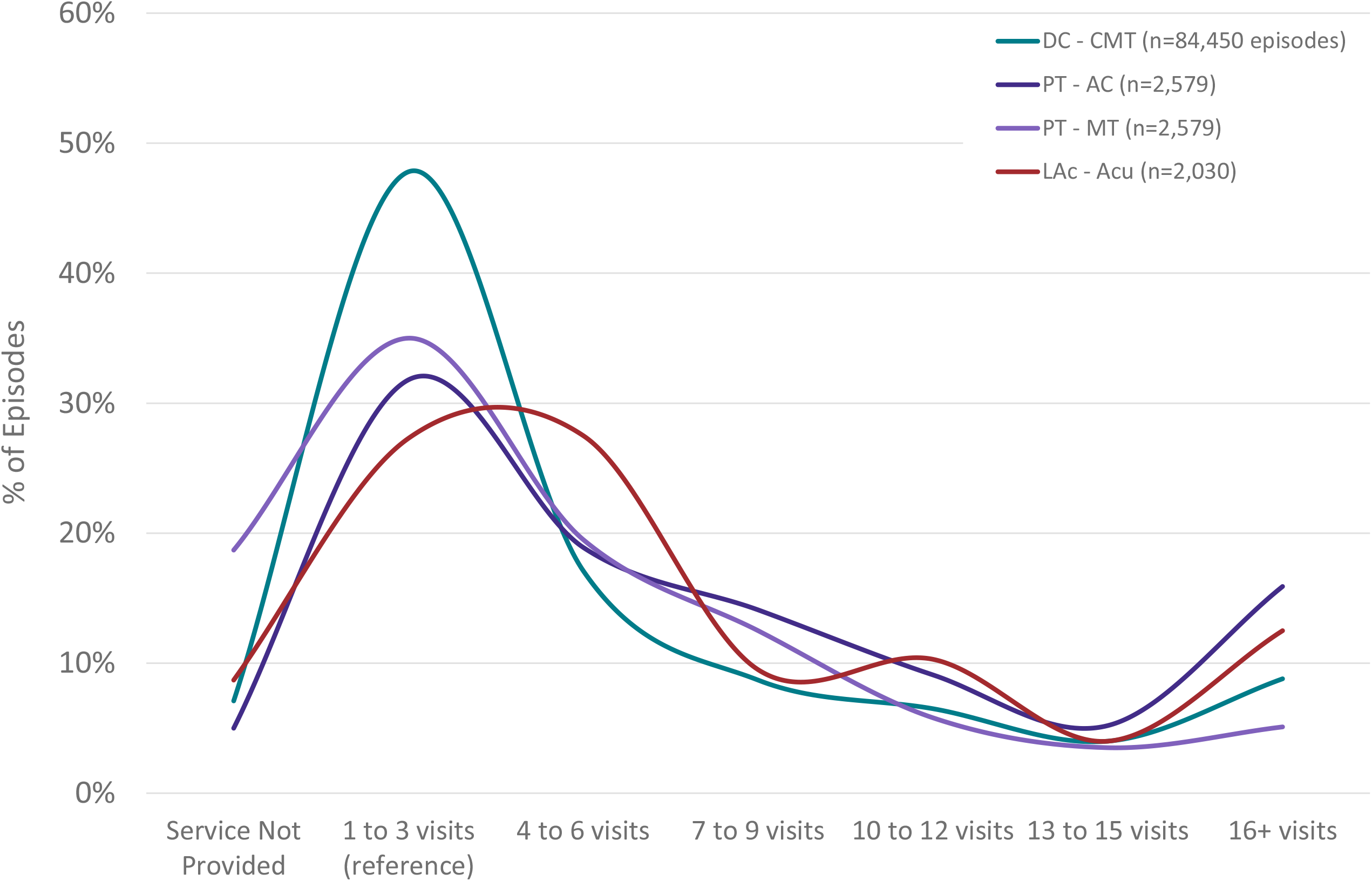
Non-surgical single episode neck pain episode distribution by type of health care provider initially contacted and # of visits of specific type of service. DC=Doctor of Chiropractic, PT=Physical Therapist, LAc=Licensed Acupuncturist, CMT=Chiropractic Manipulative Treatment, AC=Active Care, MT=Manual Therapy, Acu=Acupuncture

Relatively small sample sizes resulted in the bivariate and RR analyses revealing an absence of statistically significant or clinically meaningful differences in the exposure to second- or third-line services when AC, MT, or acupuncture were not provided, or when greater than 3 visits were provided. When CMT was not provided by a DC, or when greater than 3 visits were provided, exposure to all second- and third-line services was higher than the 1 to 3 visit reference category. (Table 1) (Table 1a). Figure 2 illustrates the RR for exposure to prescription opioids.

**Table 1.** % of single episode non-surgical neck pain episodes including specific services by type of initial contact health care provider

|  |  |  |  |  |  |  |  |  |  |
| --- | --- | --- | --- | --- | --- | --- | --- | --- | --- |
| % of Episodes Including |  | Service Not Provided | 1 to 3 visits (reference) | 4 to 6 visits | 7 to 9 visits | 10 to 12 visits | 13 to 15 visits | 16+ visits | Total |
| Initial Contact With Chiropractor (DC) - # of Visits of Chiropractic Manipulative Treatment (CMT) |  |  |  |  |  |  |  |  |  |
| First Line | Chiropractic Manipulation | 0.0% | 100.0% | 100.0% | 100.0% | 100.0% | 100.0% | 100.0% | 92.9% |
|  | Active Care | 42.6% | 23.7% | 31.7% | 35.5% | 37.1% | 37.7% | 45.0% | 30.8% |
|  | Manual Therapy | 21.5% | 16.2% | 20.3% | 21.8% | 22.2% | 22.8% | 26.6% | 19.3% |
|  | Acupuncture | 1.0% | 0.5% | 0.8% | 1.2% | 0.9% | 0.9% | 1.6% | 0.8% |
|  | Passive Therapy | 16.7% | 31.6% | 39.6% | 43.0% | 43.7% | 47.0% | 51.9% | 36.1% |
|  | Osteopathic Manipulation | 0.5% | 0.1% | 0.2% | 0.2% | 0.1% | 0.2% | 0.2% | 0.2% |
| Second Line | Imaging - Radiography | 38.6% | 12.8% | 20.1% | 26.2% | 30.1% | 32.5% | 37.0% | 21.1% |
|  | Rx - NSAID | 4.7% | 3.6% | 4.8% | 5.6% | 5.8% | 6.3% | 7.5% | 4.7% |
|  | Rx - Skeletal Muscle Relaxant | 6.3% | 2.9% | 3.7% | 4.2% | 4.0% | 5.3% | 5.6% | 3.8% |
|  | Imaging - MRI | 2.8% | 1.3% | 2.2% | 3.0% | 3.2% | 3.5% | 4.0% | 2.2% |
| Third Line | Rx - Opioid | 3.4% | 2.5% | 3.1% | 3.8% | 4.3% | 4.5% | 5.5% | 3.2% |
|  | Spinal Injection | 0.9% | 0.4% | 0.8% | 1.0% | 1.5% | 1.5% | 2.4% | 0.8% |
|  | Imaging - CT | 0.9% | 0.5% | 0.7% | 0.8% | 0.6% | 0.8% | 0.8% | 0.6% |
| Initial Contact With Physical Therapist (PT) - # of Visits of Active Care (AC) |  |  |  |  |  |  |  |  |  |
| First Line | Chiropractic Manipulation | 13.3% | 8.6% | 8.6% | 10.7% | 9.8% | 12.6% | 16.6% | 10.7% |
|  | Active Care | 0.0% | 100.0% | 100.0% | 100.0% | 100.0% | 100.0% | 100.0% | 95.0% |
|  | Manual Therapy | 66.4% | 71.0% | 85.7% | 85.7% | 88.0% | 94.8% | 89.5% | 81.3% |
|  | Acupuncture | 5.5% | 2.1% | 1.8% | 1.7% | 0.4% | 0.7% | 2.2% | 1.9% |
|  | Passive Therapy | 28.1% | 22.9% | 30.5% | 37.2% | 34.6% | 41.5% | 43.2% | 31.9% |
|  | Osteopathic Manipulation | 1.6% | 0.6% | 0.6% | 0.8% | 1.7% | 0.0% | 0.7% | 0.8% |
| Second Line | Imaging - Radiography | 26.6% | 20.5% | 20.5% | 22.3% | 19.2% | 24.4% | 26.3% | 22.1% |
|  | Rx - NSAID | 12.5% | 8.0% | 8.8% | 11.8% | 10.3% | 11.1% | 14.9% | 10.4% |
|  | Rx - Skeletal Muscle Relaxant | 14.8% | 7.6% | 8.4% | 12.4% | 2.1% | 13.3% | 12.0% | 9.3% |
|  | Imaging - MRI | 14.8% | 14.0% | 15.2% | 19.8% | 16.7% | 18.5% | 21.5% | 16.8% |
| Third Line | Rx - Opioid | 5.5% | 5.2% | 4.9% | 7.7% | 3.8% | 5.2% | 7.6% | 5.8% |
|  | Spinal Injection | 1.6% | 2.9% | 2.7% | 3.6% | 4.7% | 4.4% | 5.1% | 3.5% |
|  | Imaging - CT | 4.7% | 2.9% | 2.0% | 2.8% | 1.7% | 5.2% | 4.4% | 3.1% |
| Initial Contact With Physical Therapist (PT) - # of Visits of Manual Therapy (MT) |  |  |  |  |  |  |  |  |  |
| First Line | Chiropractic Manipulation | 10.4% | 9.0% | 9.4% | 11.1% | 13.4% | 12.2% | 24.4% | 10.7% |
|  | Active Care | 91.1% | 93.0% | 97.8% | 97.5% | 98.7% | 100.0% | 99.2% | 95.0% |
|  | Manual Therapy | 0.0% | 100.0% | 100.0% | 100.0% | 100.0% | 100.0% | 100.0% | 81.3% |
|  | Acupuncture | 2.3% | 1.7% | 1.8% | 1.5% | 1.3% | 4.4% | 3.1% | 1.9% |
|  | Passive Therapy | 17.5% | 26.7% | 35.7% | 44.3% | 42.3% | 44.4% | 55.0% | 31.9% |
|  | Osteopathic Manipulation | 0.6% | 1.1% | 0.2% | 1.2% | 0.0% | 2.2% | 0.0% | 0.8% |
| Second Line | Imaging - Radiography | 23.5% | 20.5% | 19.7% | 21.4% | 24.8% | 25.6% | 32.8% | 22.1% |
|  | Rx - NSAID | 10.2% | 8.2% | 10.4% | 12.4% | 12.8% | 11.1% | 18.3% | 10.4% |
|  | Rx - Skeletal Muscle Relaxant | 6.9% | 8.9% | 8.4% | 8.0% | 15.4% | 13.3% | 17.6% | 9.3% |
|  | Imaging - MRI | 16.2% | 13.6% | 15.5% | 17.6% | 23.5% | 23.3% | 30.5% | 16.8% |
| Third Line | Rx - Opioid | 4.6% | 5.1% | 6.2% | 5.0% | 5.4% | 8.9% | 13.7% | 5.8% |
|  | Spinal Injection | 1.2% | 2.7% | 3.6% | 5.9% | 4.7% | 6.7% | 7.6% | 3.5% |
|  | Imaging - CT | 4.6% | 2.5% | 1.4% | 4.0% | 0.7% | 6.7% | 5.3% | 3.1% |
| Initial Contact with Licensed Acupuncturist (LAc) - # of Visits of Acupuncture (Acu) |  |  |  |  |  |  |  |  |  |
| First Line | Chiropractic Manipulation | 27.7% | 9.4% | 7.3% | 9.3% | 7.6% | 12.2% | 18.9% | 11.5% |
|  | Active Care | 32.2% | 11.7% | 13.6% | 12.4% | 17.1% | 25.6% | 24.8% | 16.8% |
|  | Manual Therapy | 68.4% | 38.2% | 38.3% | 41.5% | 43.8% | 39.0% | 55.1% | 43.9% |
|  | Acupuncture | 0.0% | 100.0% | 100.0% | 100.0% | 100.0% | 100.0% | 100.0% | 91.3% |
|  | Passive Therapy | 54.2% | 33.0% | 41.1% | 39.4% | 37.6% | 51.2% | 47.6% | 40.7% |
|  | Osteopathic Manipulation | 1.7% | 0.7% | 0.2% | 0.5% | 0.5% | 0.0% | 1.6% | 0.7% |
| Second Line | Imaging - Radiography | 8.5% | 4.3% | 3.9% | 7.8% | 3.3% | 4.9% | 8.3% | 5.3% |
|  | Rx - NSAID | 4.5% | 2.7% | 2.1% | 3.6% | 2.9% | 3.7% | 3.5% | 3.0% |
|  | Rx - Skeletal Muscle Relaxant | 6.2% | 2.3% | 1.4% | 2.6% | 2.4% | 1.2% | 2.4% | 2.4% |
|  | Imaging - MRI | 4.0% | 2.3% | 1.6% | 2.6% | 3.3% | 4.9% | 7.5% | 3.2% |
| Third Line | Rx - Opioid | 3.4% | 4.3% | 2.3% | 2.6% | 2.9% | 1.2% | 3.1% | 3.1% |
|  | Spinal Injection | 2.3% | 0.7% | 0.9% | 1.0% | 1.9% | 4.9% | 2.4% | 1.4% |
|  | Imaging - CT | 0.6% | 0.4% | 0.2% | 0.0% | 0.5% | 0.0% | 0.4% | 0.3% |

Cells with red text denote that the effect of number of visits on service usage was found not to be significantly different from that of 1-3 visit reference (Fisher's Exact p > 0.001)
Cells with black text denote that the effect of number of visits on service usage was found to be significantly different from that of 1-3 visit reference (Fisher's Exact p < 0.001)

**Table 1a.** Risk ratio and 95% confidence interval for comparing % of single episode non-surgical neck pain episodes including specific services by type of initial contact health care provider and number of visits of select first line service to 1 to 3 visit reference

|  |  |  |  |  |  |  |  |  |
| --- | --- | --- | --- | --- | --- | --- | --- | --- |
| Risk ratio and 95% confidence interval |  | Service Not Provided | 1 to 3 visits | 4 to 6 visits | 7 to 9 visits | 10 to 12 visits | 13 to 15 visits | 16+ visits |
| Initial Contact With Chiropractor (DC) - # of Visits of Chiropractic Manipulative Treatment (CMT) |  |  |  |  |  |  |  |  |
| First Line | Chiropractic Manipulation | N/A | reference | 1.00 (1.00, 1.00) | 1.00 (1.00, 1.00) | 1.00 (1.00, 1.00) | 1.00 (1.00, 1.00) | 1.00 (1.00, 1.00) |
|  | Active Care | 1.80 (1.74, 1.86) |  | 1.34 (1.30, 1.38) | 1.50 (1.44, 1.55) | 1.57 (1.51, 1.63) | 1.59 (1.52, 1.67) | 1.90 (1.84, 1.96) |
|  | Manual Therapy | 1.33 (1.26, 1.40) |  | 1.25 (1.21, 1.31) | 1.35 (1.28, 1.41) | 1.37 (1.30, 1.45) | 1.41 (1.32, 1.50) | 1.64 (1.57, 1.72) |
|  | Acupuncture | 2.00 (1.50, 2.66) |  | 1.56 (1.24, 1.97) | 2.37 (1.84, 3.04) | 1.87 (1.37, 2.53) | 1.73 (1.17, 2.54) | 3.11 (2.48, 3.90) |
|  | Passive Therapy | 0.53 (0.50, 0.56) |  | 1.26 (1.22, 1.29) | 1.36 (1.32, 1.40) | 1.38 (1.34, 1.43) | 1.49 (1.43, 1.55) | 1.64 (1.60, 1.69) |
|  | Osteopathic Manipulation | 5.90 (3.59, 9.71) |  | 1.95 (1.14, 3.32) | 2.83 (1.58, 5.08) | 1.57 (0.69, 3.54) | 2.19 (0.92, 5.21) | 2.13 (1.12, 4.05) |
| Second Line | Imaging - Radiography | 3.02 (2.90, 3.14) |  | 1.57 (1.51, 1.64) | 2.05 (1.95, 2.14) | 2.35 (2.24, 2.47) | 2.54 (2.40, 2.68) | 2.90 (2.79, 3.01) |
|  | Rx - NSAID | 1.31 (1.16, 1.48) |  | 1.32 (1.21, 1.44) | 1.55 (1.39, 1.72) | 1.60 (1.42, 1.80) | 1.75 (1.53, 2.02) | 2.08 (1.90, 2.29) |
|  | Rx - Skeletal Muscle Relaxant | 2.14 (1.92, 2.40) |  | 1.27 (1.15, 1.40) | 1.43 (1.27, 1.62) | 1.36 (1.18, 1.56) | 1.79 (1.54, 2.09) | 1.92 (1.72, 2.14) |
|  | Imaging - MRI | 2.10 (1.77, 2.49) |  | 1.71 (1.49, 1.96) | 2.26 (1.93, 2.64) | 2.42 (2.05, 2.87) | 2.63 (2.16, 3.20) | 3.01 (2.62, 3.46) |
| Third Line | Rx-Opioid | 1.40 (1.21, 1.62) |  | 1.26 (1.13, 1.41) | 1.55 (1.36, 1.76) | 1.74 (1.51, 2.00) | 1.85 (1.56, 2.18) | 2.24 (2.00, 2.51) |
|  | Spinal Injection | 2.45 (1.81, 3.32) |  | 2.08 (1.64, 2.64) | 2.68 (2.04, 3.52) | 3.79 (2.90, 4.96) | 3.85 (2.81, 5.29) | 6.11 (4.93, 7.57) |
|  | Imaging-CT | 1.97 (1.47, 2.65) |  | 1.40 (1.10, 1.78) | 1.76 (1.32, 2.33) | 1.22 (0.84, 1.77) | 1.74 (1.17, 2.58) | 1.76 (1.33, 2.33) |
| Initial Contact With Physical Therapist (PT) - # of Visits of Active Care (AC) |  |  |  |  |  |  |  |  |
| First Line | Chiropractic Manipulation | 1.54 (0.94, 2.52) | reference | 1.00 (0.69, 1.43) | 1.24 (0.86, 1.80) | 1.14 (0.73, 1.78) | 1.46 (0.89, 2.39) | 1.92 (1.41, 2.62) |
|  | Active Care | N/A |  | 1.00 (1.00, 1.00) | 1.00 (1.00, 1.00) | 1.00 (1.00, 1.00) | 1.00 (1.00, 1.00) | 1.00 (1.00, 1.00) |
|  | Manual Therapy | 0.94 (0.82, 1.07) |  | 1.21 (1.14, 1.28) | 1.21 (1.14, 1.28) | 1.24 (1.16, 1.32) | 1.34 (1.26, 1.42) | 1.26 (1.19, 1.33) |
|  | Acupuncture | 2.64 (1.12, 6.24) |  | 0.89 (0.40, 1.98) | 0.80 (0.32, 2.01) | 0.21 (0.03, 1.54) | 0.36 (0.05, 2.67) | 1.06 (0.48, 2.36) |
|  | Passive Therapy | 1.23 (0.91, 1.66) |  | 1.33 (1.11, 1.60) | 1.62 (1.35, 1.95) | 1.51 (1.22, 1.88) | 1.81 (1.43, 2.29) | 1.89 (1.59, 2.23) |
|  | Osteopathic Manipulation | 2.57 (0.50, 13.08) |  | 1.01 (0.24, 4.21) | 1.36 (0.33, 5.65) | 2.81 (0.76, 10.37) | N/A | 1.20 (0.29, 5.00) |
| Second Line | Imaging - Radiography | 1.30 (0.94, 1.78) |  | 1.00 (0.80, 1.25) | 1.09 (0.86, 1.38) | 0.94 (0.70, 1.26) | 1.19 (0.86, 1.65) | 1.29 (1.04, 1.59) |
|  | Rx - NSAID | 1.55 (0.93, 2.60) |  | 1.10 (0.76, 1.58) | 1.47 (1.02, 2.12) | 1.28 (0.82, 1.99) | 1.38 (0.81, 2.35) | 1.85 (1.33, 2.57) |
|  | Rx - Skeletal Muscle Relaxant | 1.97 (1.22, 3.17) |  | 1.11 (0.76, 1.62) | 1.64 (1.14, 2.36) | 0.28 (0.12, 0.70) | 1.77 (1.08, 2.89) | 1.58 (1.11, 2.26) |
|  | Imaging - MRI | 1.06 (0.68, 1.66) |  | 1.08 (0.83, 1.42) | 1.42 (1.08, 1.85) | 1.19 (0.85, 1.66) | 1.32 (0.89, 1.96) | 1.53 (1.19, 1.97) |
| Third Line | Rx-Opioid | 1.04 (0.48, 2.27) |  | 0.94 (0.58, 1.53) | 1.47 (0.93, 2.33) | 0.73 (0.36, 1.48) | 0.99 (0.45, 2.15) | 1.44 (0.92, 2.26) |
|  | Spinal Injection | 0.53 (0.13, 2.23) |  | 0.91 (0.47, 1.77) | 1.23 (0.63, 2.38) | 1.61 (0.80, 3.23) | 1.52 (0.63, 3.65) | 1.75 (0.99, 3.11) |
|  | Imaging-CT | 1.60 (0.67, 3.85) |  | 0.70 (0.34, 1.45) | 0.94 (0.46, 1.95) | 0.58 (0.20, 1.67) | 1.77 (0.78, 4.04) | 1.50 (0.82, 2.74) |
| Initial Contact With Physical Therapist (PT) - # of Visits of Manual Therapy (MT) |  |  |  |  |  |  |  |  |
| First Line | Chiropractic Manipulation | 1.16 (0.83, 1.62) | reference | 1.04 (0.74, 1.47) | 1.24 (0.86, 1.80) | 1.50 (0.95, 2.36) | 1.36 (0.75, 2.46) | 2.72 (1.89, 3.93) |
|  | Active Care | 0.98 (0.95, 1.01) |  | 1.05 (1.03, 1.07) | 1.05 (1.02, 1.07) | 1.06 (1.03, 1.09) | 1.07 (1.06, 1.09) | 1.07 (1.04, 1.09) |
|  | Manual Therapy | N/A |  | 1.00 (1.00, 1.00) | 1.00 (1.00, 1.00) | 1.00 (1.00, 1.00) | 1.00 (1.00, 1.00) | 1.00 (1.00, 1.00) |
|  | Acupuncture | 1.38 (0.64, 2.97) |  | 1.08 (0.48, 2.45) | 0.93 (0.34, 2.54) | 0.81 (0.19, 3.50) | 2.68 (0.91, 7.89) | 1.84 (0.62, 5.45) |
|  | Passive Therapy | 0.65 (0.52, 0.82) |  | 1.34 (1.14, 1.57) | 1.66 (1.41, 1.95) | 1.58 (1.28, 1.97) | 1.67 (1.29, 2.15) | 2.06 (1.70, 2.49) |
|  | Osteopathic Manipulation | 0.56 (0.16, 2.04) |  | 0.18 (0.02, 1.40) | 1.12 (0.35, 3.54) | N/A | 2.01 (0.45, 9.02) | N/A |
| Second Line | Imaging - Radiography | 1.15 (0.93, 1.41) |  | 0.96 (0.77, 1.20) | 1.04 (0.82, 1.33) | 1.21 (0.89, 1.65) | 1.25 (0.86, 1.82) | 1.60 (1.21, 2.11) |
|  | Rx - NSAID | 1.24 (0.88, 1.75) |  | 1.26 (0.90, 1.77) | 1.51 (1.05, 2.17) | 1.56 (0.97, 2.50) | 1.36 (0.73, 2.53) | 2.24 (1.47, 3.41) |
|  | Rx - Skeletal Muscle Relaxant | 0.77 (0.52, 1.14) |  | 0.94 (0.66, 1.35) | 0.91 (0.59, 1.39) | 1.74 (1.13, 2.68) | 1.50 (0.85, 2.65) | 1.98 (1.29, 3.03) |
|  | Imaging - MRI | 1.19 (0.92, 1.55) |  | 1.14 (0.88, 1.48) | 1.30 (0.97, 1.73) | 1.72 (1.24, 2.41) | 1.71 (1.14, 2.58) | 2.24 (1.65, 3.04) |
| Third Line | Rx-Opioid | 0.90 (0.55, 1.47) |  | 1.21 (0.78, 1.89) | 0.97 (0.56, 1.69) | 1.05 (0.51, 2.19) | 1.74 (0.85, 3.58) | 2.70 (1.61, 4.51) |
|  | Spinal Injection | 0.47 (0.19, 1.14) |  | 1.35 (0.74, 2.46) | 2.21 (1.23, 3.99) | 1.77 (0.78, 4.03) | 2.51 (1.05, 5.98) | 2.87 (1.41, 5.87) |
|  | Imaging-CT | 1.80 (1.01, 3.19) |  | 0.55 (0.24, 1.27) | 1.58 (0.81, 3.08) | 0.26 (0.04, 1.94) | 2.62 (1.09, 6.26) | 2.10 (0.92, 4.79) |
| Initial Contact with Licensed Acupuncturist (LAc) - # of Visits of Acupuncture (Acu) |  |  |  |  |  |  |  |  |
| First Line | Chiropractic Manipulation | 2.95 (2.08, 4.20) | reference | 0.78 (0.53, 1.16) | 1.00 (0.60, 1.66) | 0.81 (0.48, 1.39) | 1.30 (0.69, 2.46) | 2.02 (1.40, 2.90) |
|  | Active Care | 2.75 (2.01, 3.76) |  | 1.16 (0.85, 1.58) | 1.06 (0.68, 1.65) | 1.46 (1.01, 2.13) | 2.19 (1.42, 3.37) | 2.12 (1.55, 2.90) |
|  | Manual Therapy | 1.79 (1.55, 2.07) |  | 1.00 (0.86, 1.16) | 1.09 (0.89, 1.32) | 1.15 (0.95, 1.38) | 1.02 (0.76, 1.37) | 1.44 (1.24, 1.68) |
|  | Acupuncture | N/A |  | 1.00 (1.00, 1.00) | 1.00 (1.00, 1.00) | 1.00 (1.00, 1.00) | 1.00 (1.00, 1.00) | 1.00 (1.00, 1.00) |
|  | Passive Therapy | 1.64 (1.37, 1.97) |  | 1.25 (1.07, 1.46) | 1.19 (0.97, 1.48) | 1.14 (0.92, 1.41) | 1.55 (1.22, 1.98) | 1.44 (1.21, 1.72) |
|  | Osteopathic Manipulation | 2.35 (0.53, 10.41) |  | 0.25 (0.03, 2.21) | 0.72 (0.08, 6.39) | 0.66 (0.07, 5.88) | N/A | 2.19 (0.55, 8.67) |
| Second Line | Imaging - Radiography | 1.96 (1.05, 3.65) |  | 0.91 (0.52, 1.60) | 1.80 (0.96, 3.35) | 0.77 (0.34, 1.76) | 1.13 (0.40, 3.17) | 1.91 (1.09, 3.37) |
|  | Rx - NSAID | 1.67 (0.72, 3.88) |  | 0.79 (0.38, 1.68) | 1.34 (0.56, 3.24) | 1.06 (0.42, 2.69) | 1.35 (0.40, 4.57) | 1.31 (0.58, 2.96) |
|  | Rx - Skeletal Muscle Relaxant | 2.65 (1.21, 5.82) |  | 0.61 (0.26, 1.46) | 1.11 (0.40, 3.06) | 1.02 (0.37, 2.82) | 0.52 (0.07, 3.93) | 1.01 (0.39, 2.62) |
|  | Imaging - MRI | 1.69 (0.68, 4.17) |  | 0.69 (0.30, 1.59) | 1.11 (0.40, 3.06) | 1.42 (0.58, 3.52) | 2.08 (0.70, 6.23) | 3.19 (1.60, 6.36) |
| Third Line | Rx-Opioid | 0.78 (0.33, 1.89) |  | 0.54 (0.28, 1.05) | 0.60 (0.23, 1.55) | 0.66 (0.27, 1.59) | 0.28 (0.04, 2.06) | 0.73 (0.33, 1.60) |
|  | Spinal Injection | 3.14 (0.79, 12.41) |  | 1.24 (0.34, 4.60) | 1.44 (0.27, 7.79) | 2.64 (0.67, 10.47) | 6.77 (1.73, 26.54) | 3.28 (0.93, 11.51) |
|  | Imaging-CT | 1.57 (0.14, 17.19) |  | 0.50 (0.05, 5.46) | N/A | 1.32 (0.12, 14.50) | N/A | 1.09 (0.10, 11.99) |

Cells with red text denote 95% confidence interval includes 1 Cells with black text denote 95% confidence interval does not include 1

**Figure 2.**
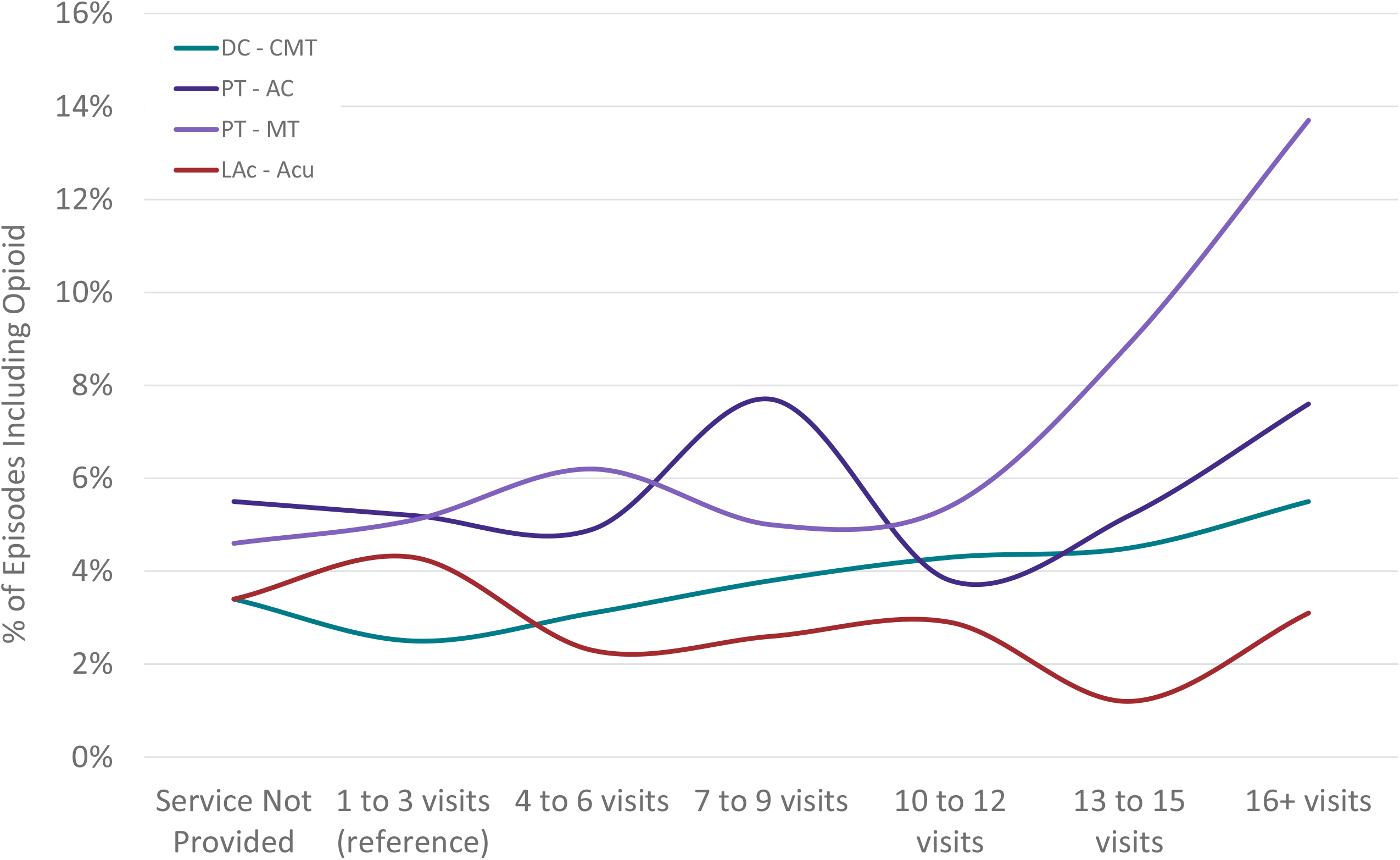
Non-surgical single episode neck pain episodes including a prescription opioid by type of health care provider initially contacted and # of visits of specific type of service. DC=Doctor of Chiropractic, PT=Physical Therapist, LAc=Licensed Acupuncturist, CMT=Chiropractic Manipulative Treatment, AC=Active Care, MT=Manual Therapy, Acu=Acupuncture

Total episode cost and episode duration increased significantly with an increasing number of visits for all HCP and service type combinations. The DC-CMT combination was associated with the lowest overall median total episode cost ($185, Q1 80, Q3 455) a finding that remained consistent within each visit count category (Table 2) (Figure 4). The median number of different HCPs seen during an episode increased with an increasing number of visits for the PT-AC and PT-MT combinations and was unchanged for the DC-CMT and LAc-acupuncture combinations.

**Table 2.** Single episode non-surgical neck pain episode characteristics by type of initial contact health care provider (HCP) and number of visits of select services

Cells with red text denote 95% confidence interval includes 1 Cells with black text denote 95% confidence interval does not include 1
|  |  |  |  |  |  |  |  |  |
| --- | --- | --- | --- | --- | --- | --- | --- | --- |
| % or Median (Q1, Q3) | Service Not Provided | 1 to 3 visits (reference) | 4 to 6 visits | 7 to 9 visits | 10 to 12 visits | 13 to 15 visits | 16+ visits | Total |
| Initial Contact With Chiropractor (DC) - # of Visits of Chiropractic Manipulative Treatment (CMT) |  |  |  |  |  |  |  |  |
| Total Cost | 1967738 | 6687048 | 5285162 | 4004025 | 3940956 | 2869185 | 9980810 | 34734923 |
| % Total Cost | 5.7% | 19.3% | 15.2% | 11.5% | 11.3% | 8.3% | 28.7% | 100.0% |
| Episode Cost | \$89 (48, 400) | \$92 (55, 146) | \$250 (195, 330) | \$420 (333, 520) | \$550 (450, 705) | \$700 (582, 844) | \$1042 (776, 1430) | \$185 (80, 455) |
| # of HCP Seen | 1 (1, 2) | 1 (1, 1) | 1 (1, 2) | 1 (1, 2) | 1 (1, 2) | 1 (1, 2) | 2 (1, 3) | 1 (1, 2) |
| Episode Duration - days | 8 (1, 39) | 5 (1, 25) | 40 (18, 102) | 77 (38, 179) | 102 (48, 247) | 148 (69, 311) | 217 (100, 353) | 29 (4, 104) |
| Initial Contact With Physical Therapist (PT) - # of Visits of Active Care (AC) |  |  |  |  |  |  |  |  |
| Total Cost | 77628 | 521588 | 432078 | 429101 | 337555 | 195573 | 877425 | 2870949 |
| % Total Cost | 2.7% | 18.2% | 15.1% | 14.9% | 11.8% | 6.8% | 30.6% | 100.0% |
| Episode Cost | \$307 (172, 735) | \$323 (165, 636) | \$528 (315, 965) | \$685 (489, 1356) | \$812 (541, 1381) | \$1061 (656, 1785) | \$1517 (971, 2542) | \$613 (322, 1309) |
| # of HCP Seen | 2 (2, 3) | 2 (1, 3) | 2 (1, 3) | 2 (1, 4) | 3 (1, 4) | 3 (2, 4) | 3 (2, 5) | 2 (1, 4) |
| Episode Duration - days | 48 (14, 135) | 23 (6, 59) | 41 (22, 85) | 50 (28, 112) | 52 (36, 103) | 77 (43, 150) | 99 (55, 233) | 46 (22, 108) |
| Initial Contact With Physical Therapist (PT) - # of Visits of Manual Therapy (MT) |  |  |  |  |  |  |  |  |
| Total Cost | 357777 | 602730 | 577803 | 442830 | 267823 | 184322 | 437663 | 2870949 |
| % Total Cost | 12.5% | 21.0% | 20.1% | 15.4% | 9.3% | 6.4% | 15.2% | 100.0% |
| Episode Cost | \$427 (190, 860) | \$369 (206, 730) | \$618 (438, 1169) | \$924 (609, 1440) | \$1216 (845, 1918) | \$1693 (1260, 2501) | \$2648 (1875, 3889) | \$613 (322, 1309) |
| # of HCP Seen | 2 (1, 3) | 2 (1, 3) | 2 (1, 4) | 3 (1, 4) | 3 (2, 5) | 4 (2, 5) | 4 (2, 7) | 2 (1, 4) |
| Episode Duration - days | 33 (10, 87) | 29 (10, 70) | 48 (29, 92) | 55 (36, 112) | 83 (48, 218) | 99 (74, 232) | 194 (102, 324) | 46 (22, 108) |
| Initial Contact with Licensed Acupuncturist (LAc) - # of Visits of Acupuncture (Acu) |  |  |  |  |  |  |  |  |
| Total Cost | 72627 | 153586 | 228937 | 116436 | 182107 | 98415 | 509782 | 1361890 |
| % Total Cost | 5.3% | 11.3% | 16.8% | 8.5% | 13.4% | 7.2% | 37.4% | 100.0% |
| Episode Cost | \$170 (59, 396) | \$133 (100, 182) | \$296 (216, 389) | \$473 (363, 574) | \$656 (487, 798) | \$868 (632, 1163) | \$1406 (1012, 2353) | \$336 (158, 715) |
| # of HCP Seen | 1 (1, 2) | 1 (1, 1) | 1 (1, 1) | 1 (1, 2) | 1 (1, 2) | 1 (1, 2) | 1 (1, 3) | 1 (1, 2) |
| Episode Duration - days | 26 (1, 64) | 1 (1, 15) | 21 (10, 42) | 40 (23, 71) | 44 (30, 100) | 51 (43, 97) | 92 (46, 226) | 29 (6, 69) |

Cells with red text denote that the effect of provider type on service usage was found not to be significantly different from that of PCP-reference (Mann-Whitney U p > 0.001)
Cells with black text denote that the effect of provider type on service usage was found to be significantly different from that of PCP-reference (Mann-Whitney U p < 0.001)

**Figure 3.**
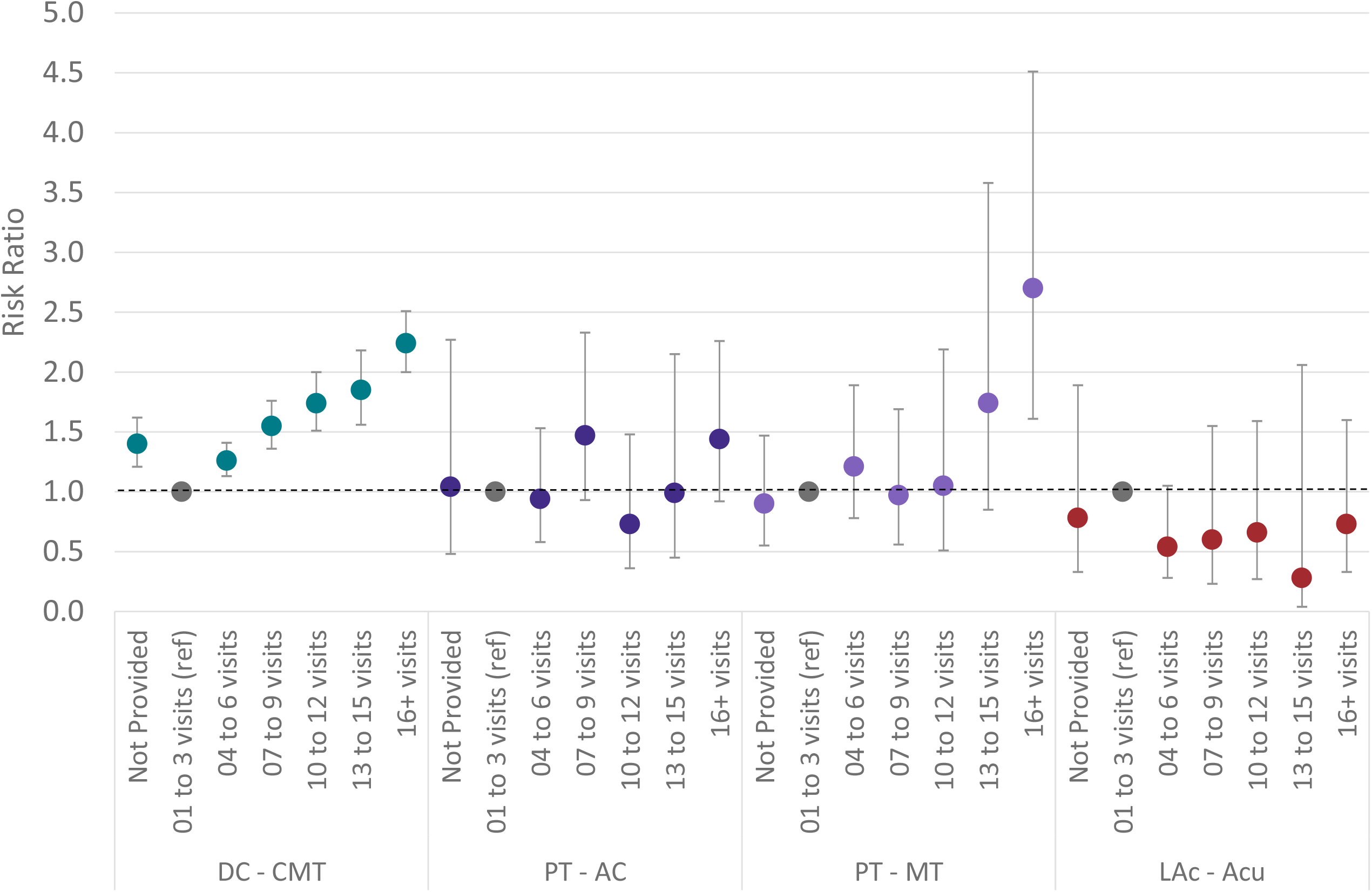
Non-surgical single episode neck pain risk ratio and 95% confidence interval for opioid exposure compared to the 1 to 3 visit reference by type of health care provider initially contacted and # of visits of specific type of service. DC=Doctor of Chiropractic, PT=Physical Therapist, LAc=Licensed Acupuncturist, CMT=Chiropractic Manipulative Treatment, AC=Active Care, MT=Manual Therapy, Acu=Acupuncture

**Figure 4.**
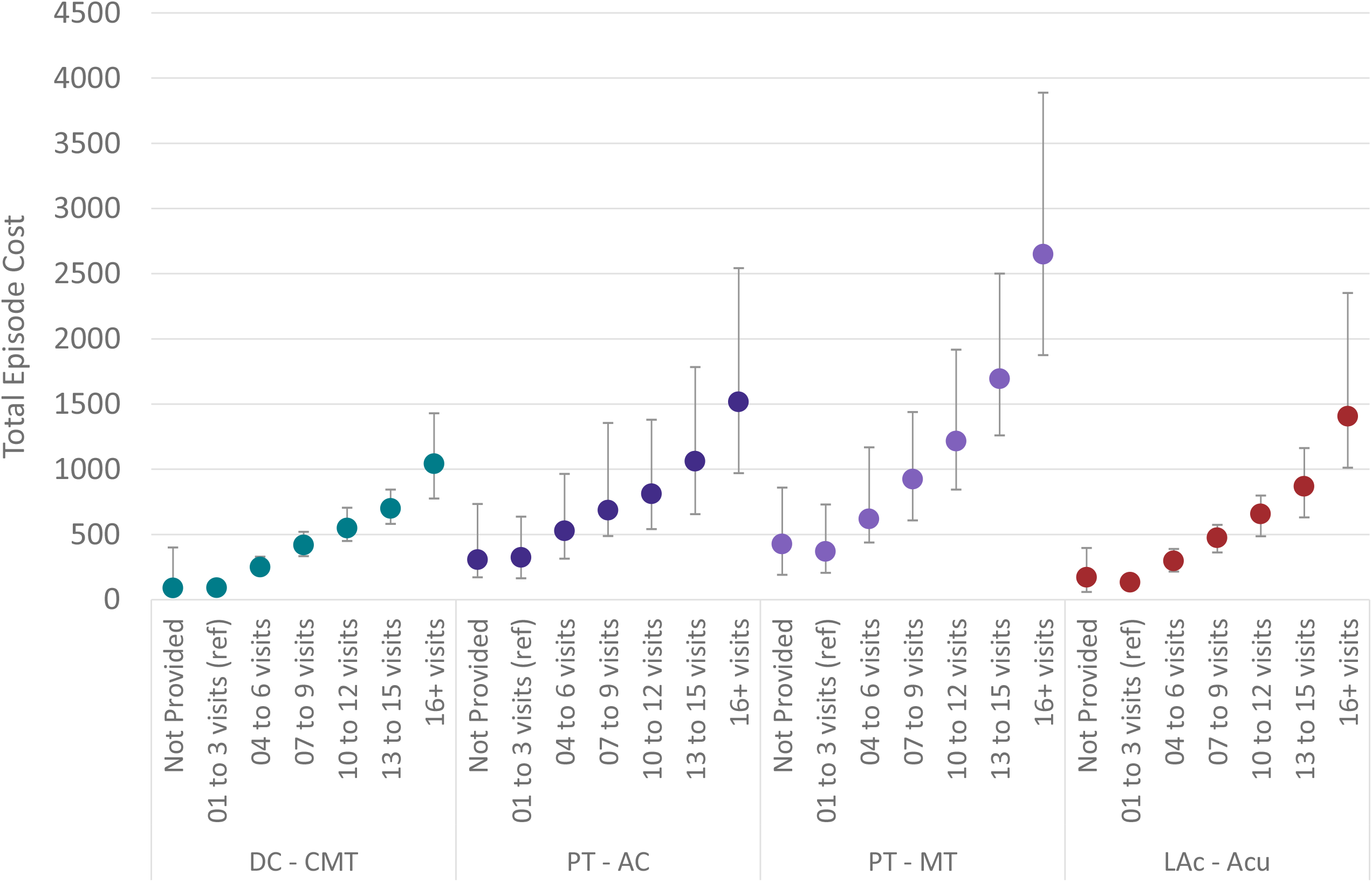
Non-surgical single episode neck pain median and interquartile range (Q1, Q3) total episode cost by type of health care provider initially contacted and # of visits of specific type of service. DC=Doctor of Chiropractic, PT=Physical Therapist, LAc=Licensed Acupuncturist, CMT=Chiropractic Manipulative Treatment, AC=Active Care, MT=Manual Therapy, Acu=Acupuncture

## Discussion

This study provides a comprehensive analysis of the relationship between the number of visits of CMT, AC, MT, and acupuncture, utilization of other health care services, and total episode cost for individuals with non-surgical NP initially contacting a DC, PT, or LAc. Like LBP ^27^, for NP the most common level of utilization of CMT, AC, MT was 1 to 3 visits, and 4 to 6 visits of acupuncture. CMT was associated with lowest median total episode cost compared to AC, MT, and acupuncture. While higher levels of visit utilization were not associated with lower rates of imaging, pharmaceutical or interventional services the study was not able to evaluate potential clinical benefits associated with different numbers of visits of CMT, AC, MT, and acupuncture.

Like any retrospective study involving administrative data this study has several limitations. Regarding commercial insurance coverage, variability in benefit plan design, variability in enrollee cost-sharing responsibility, and missing information were potential sources of confounding or bias. Risk is minimized as the cohort had continuous highly uniform commercial insurance coverage and the processing of administrative claims data included extensive quality and actuarial control measures. The commercial insurer HCP database may have included errors or missing information, however the risk of misclassifying DCs, PTs and LAcs is minimized through continuous credentialing and HCP data validation processes. Variation in insurance coverage, network participation contracts, alternative reimbursement models, and individuals seeking treatment outside of the insurance benefit are associated with limitations in summarizing total episode cost. While individuals from all 50 states, the cohort did not describe a US representative sample.

Analyses involving the type of HCP initially contacted by an individual with NP or LBP must contend with the risk of confounding and bias associated with the limited ability to control for individual preference for the type of initial contact HCP and potentially meaningful differences in clinical complexity of individuals seeking services from different types of providers. These risks were minimized, but no eliminated, in this study by limiting the cohort to individuals initially contacting a DC, PT, or LAc, and by excluding NP associated with serious pathology, individuals with multiple episodes of NP, and episodes involving a surgical procedure. More important for this study was an inability to control for individual expectations or requests for specific health care services and WTP for different numbers of visits. An analysis of the change in patient functional status associated with different numbers of visits of CMT, AC, MT, and acupuncture was not possible given the absence of baseline and sequential patient reported outcome data.

This study corroborates and expands upon previous similar studies. When initially contacted by an individual with NP, DCs, PTs, and LAcs are generally associated with greater use of non-pharmaceutical and non-interventional services than primary care, specialist, and emergency medicine/urgent care HCPs.^14^ This study expands on these observed associations, demonstrating that this guideline concordance benefit is associated with as few as 1 to 3 visits of CMT, AC, MT, and acupuncture services. The study findings corroborate and are nearly identical to a similar study of dose/response of CMT, AC, MT, and acupuncture for LBP.^27^ The finding that 1-3 visits of CMT was most common for NP and that this dose level was associated with the lowest total cost corroborates an earlier study ^24^ however the earlier study’s description of this level of dose as discordant with guidelines warrants further study given the finding that this dose level is both the most common and also associated with the lowest level of utilization of pharmaceutical, imaging, and interventional services.

## Conclusions

For individuals with a single episode of non-surgical NP initially contacting a DC, PT, or LAc, 1 to 3 visits of CMT, AC, or MT, or 4 to 6 visits of acupuncture, are common and associated with avoidance of pharmaceutical, imaging, or interventional services. Not surprisingly, increasing the number of visits of CMT, AC, MT or acupuncture is associated with total episode cost, however higher numbers of visits are not associated with additional pharmaceutical, imaging, or interventional service avoidance benefits. There are a range of potential confounders to consider when interpreting or translating these findings, and there may have been unmeasured clinical benefits associated with, a higher number of visits of CMT, AC, MT, or acupuncture.

## Supporting information

Supplement - State Summary

Supplement - STROBE Checklist

Supplement 1 - Distribution of Episodes

Supplement 2 - Cohort Summary

Supplement 3 - Cohort Details

## Data Availability

All data produced in the present work are contained in the manuscript

## List of Abbreviations

LBP: Low back pain
NP: Neck pain
US: United States
CPG: Clinical practice guideline
DC: Doctor of Chiropractic
PT: Physical Therapist
LAc: Licensed Acupuncturist
HCP: Health care provider
ADI: Area Deprivation Index
NHW: Non-Hispanic white
WTP: Willingness to pay
STROBE: Strengthening the Reporting of Observational Studies in Epidemiology
ETG^*®*^: Episode Treatment Group^*®*^
ERG^*®*^: Episode Risk Group^*®*^
ACP: American College of Physicians
RR: Risk ratio
OR: Odds ratio
SD: Standard deviation
CMT: Chiropractic manipulative treatment
AC: Active care
MT: Manual therapy

## References

1. Kazeminasab S, Nejadghaderi SA, Amiri P, et al. Neck pain: global epidemiology, trends and risk factors. BMC Musculoskelet Disord. Jan 3 2022;23(1):26. doi:10.1186/s12891-021-04957-4

2. Safiri S, Kolahi AA, Hoy D, et al. Global, regional, and national burden of neck pain in the general population, 1990-2017: systematic analysis of the Global Burden of Disease Study 2017. BMJ. Mar 26 2020;368:m791. doi:10.1136/bmj.m791

3. Dieleman JL, Cao J, Chapin A, et al. US Health Care Spending by Payer and Health Condition, 1996-2016. JAMA. Mar 3 2020;323(9):863–884. doi:10.1001/jama.2020.0734

4. Qaseem A, Wilt TJ, McLean RM, et al. Noninvasive Treatments for Acute, Subacute, and Chronic Low Back Pain: A Clinical Practice Guideline From the American College of Physicians. Ann Intern Med. Apr 4 2017;166(7):514–530. doi:10.7326/M16-2367

5. Meroni R, Piscitelli D, Ravasio C, et al. Evidence for managing chronic low back pain in primary care: a review of recommendations from high-quality clinical practice guidelines. Disabil Rehabil. Apr 2021;43(7):1029–1043. doi:10.1080/09638288.2019.1645888

6. Foster NE, Anema JR, Cherkin D, et al. Prevention and treatment of low back pain: evidence, challenges, and promising directions. Lancet. Jun 9 2018;391(10137):2368–2383. doi:10.1016/S0140-6736(18)30489-6

7. Bier JD, Scholten-Peeters WGM, Staal JB, et al. Clinical Practice Guideline for Physical Therapy Assessment and Treatment in Patients With Nonspecific Neck Pain. Phys Ther. Mar 1 2018;98(3):162–171. doi:10.1093/ptj/pzx118

8. Blanpied PR, Gross AR, Elliott JM, et al. Neck Pain: Revision 2017. J Orthop Sports Phys Ther. Jul 2017;47(7):A1–A83. doi:10.2519/jospt.2017.0302

9. Cote P, Wong JJ, Sutton D, et al. Management of neck pain and associated disorders: A clinical practice guideline from the Ontario Protocol for Traffic Injury Management (OPTIMa) Collaboration. Eur Spine J. Jul 2016;25(7):2000–22. doi:10.1007/s00586-016-4467-7

10. Horn ME, Brennan GP, George SZ, Harman JS, Bishop MD. A value proposition for early physical therapist management of neck pain: a retrospective cohort analysis. BMC Health Serv Res. Jul 12 2016;16:253. doi:10.1186/s12913-016-1504-5

11. Horn ME, Fritz JM. Timing of physical therapy consultation on 1-year healthcare utilization and costs in patients seeking care for neck pain: a retrospective cohort. BMC Health Serv Res. Nov 26 2018;18(1):887. doi:10.1186/s12913-018-3699-0

12. Louis CJ, Herrera CS, Garrity BM, et al. Association of Initial Provider Type on Opioid Fills for Individuals With Neck Pain. Arch Phys Med Rehabil. Aug 2020;101(8):1407–1413. doi:10.1016/j.apmr.2020.04.002

13. Roblin DW, Liu H, Cromwell LF, et al. Provider type and management of common visits in primary care. Am J Manag Care. Apr 2017;23(4):225–231.

14. Elton D, Zhang, Meng. Neck pain care pathways and costs: association with the type of initial contact health care provider. A retrospective cohort study (preprint). medRxiv. 2022;doi:10.1101/2022.07.18.22277777v1

15. Elton D, Kosloff TM, Zhang M, et al. Low back pain care pathways and costs: association with the type of initial contact health care provider. A retrospective cohort study (preprint). medRxiv. 2022:2022.06.17.22276443. doi:10.1101/2022.06.17.22276443

16. Haas M, Groupp E, Kraemer DF. Dose-response for chiropractic care of chronic low back pain. Spine J. Sep-Oct 2004;4(5):574–83. doi:10.1016/j.spinee.2004.02.008

17. Haas M, Vavrek D, Peterson D, Polissar N, Neradilek MB. Dose-response and efficacy of spinal manipulation for care of chronic low back pain: a randomized controlled trial. Spine J. Jul 1 2014;14(7):1106–16. doi:10.1016/j.spinee.2013.07.468

18. Herman PM, Edgington SE, Sorbero ME, Hurwitz EL, Goertz CM, Coulter ID. Visit Frequency and Outcomes for Patients Using Ongoing Chiropractic Care for Chronic Low-Back and Neck Pain: An Observational Longitudinal Study. Pain Physician. Jan 2021;24(1):E61–E74.

19. Long CR, Lisi AJ, Vining RD, et al. Veteran Response to Dosage in Chiropractic Therapy (VERDICT): Study Protocol of a Pragmatic Randomized Trial for Chronic Low Back Pain. Pain Med. Dec 12 2020;21(Suppl 2):S37–S44. doi:10.1093/pm/pnaa289

20. Mueller J, Niederer D. Dose-response-relationship of stabilisation exercises in patients with chronic non-specific low back pain: a systematic review with meta-regression. Sci Rep. Oct 9 2020;10(1):16921. doi:10.1038/s41598-020-73954-9

21. Vavrek DA, Sharma R, Haas M. Cost analysis related to dose-response of spinal manipulative therapy for chronic low back pain: outcomes from a randomized controlled trial. J Manipulative Physiol Ther. Jun 2014;37(5):300–11. doi:10.1016/j.jmpt.2014.03.002

22. Pasquier M, Daneau C, Marchand AA, Lardon A, Descarreaux M. Spinal manipulation frequency and dosage effects on clinical and physiological outcomes: a scoping review. Chiropr Man Therap. 2019;27:23. doi:10.1186/s12998-019-0244-0

23. Haas M, Groupp E, Aickin M, et al. Dose response for chiropractic care of chronic cervicogenic headache and associated neck pain: a randomized pilot study. J Manipulative Physiol Ther. Nov-Dec 2004;27(9):547–53. doi:10.1016/j.jmpt.2004.10.007

24. Lewing B, Contreras J, Sansgiry SS. Demographics of and Costs to Users of Chiropractic Services in the United States with Neck or Back Pain not Meeting Guideline-Based Minimum Treatment Frequency Thresholds. Altern Ther Health Med. Oct 15 2021;

25. Schwehr NA, Shippee ND, Johnson PJ. Acupuncture ‘dose’ (number of treatments) and insurance benefits in the USA. Acupunct Med. Apr 2018;36(2):88–95. doi:10.1136/acupmed-2016-011341

26. Herman PM, Luoto JE, Kommareddi M, Sorbero ME, Coulter ID. Patient Willingness to Pay for Reductions in Chronic Low Back Pain and Chronic Neck Pain. J Pain. Nov 2019;20(11):1317–1327. doi:10.1016/j.jpain.2019.05.002

27. Elton D, Zhang M. Low back pain service utilization and costs: association with number of visits of chiropractic manipulation, active care, or manual therapy. A retrospective cohort study. medRxiv. 2022:2022.10.28.22281664. doi:10.1101/2022.10.28.22281664

28. Service IR. US Department of Treasury SOI tax stats - individual income tax statistics - 2017 ZIP code data. Accessed July 20, 2020. https://www.irs.gov/statistics/soi-tax-stats-individual-income-tax-statistics-2017-zip-code-data-soi.

29. Bureau USC. ACS Demographic and Housing Estimates. Accessed July 8, 2020. https://data.census.gov/cedsci/table?q=United%20States&g=0100000US&y=2018&tid=ACSDP1Y2018.DP05&hidePreview=true.

30. Kind AJH, Buckingham WR. Making Neighborhood-Disadvantage Metrics Accessible - The Neighborhood Atlas. N Engl J Med. Jun 28 2018;378(26):2456–2458. doi:10.1056/NEJMp1802313

31. von Elm E, Altman DG, Egger M, et al. The Strengthening the Reporting of Observational Studies in Epidemiology (STROBE) Statement: guidelines for reporting observational studies. Int J Surg. Dec 2014;12(12):1495–9. doi:10.1016/j.ijsu.2014.07.013

32. Hernan MA. The C-Word: Scientific Euphemisms Do Not Improve Causal Inference From Observational Data. Am J Public Health. May 2018;108(5):616–619. doi:10.2105/AJPH.2018.304337

33. Smith GD, Ebrahim S. Data dredging, bias, or confounding. BMJ. Dec 21 2002;325(7378):1437–8. doi:10.1136/bmj.325.7378.1437

34. King G NR. Why Propensity Scores Should Not Be Used for Matching. Political Analysis. 2019;27(4):435–454. doi:10.1017/pan.2019.11

35. National Academies of Sciences E, and Medicine. Health-Care Utilization as a Proxy in Disability Determination. 2 Factors That Affect Health-Care Utilization. National Academies Press 2018.

36. Insight O. Symmetry episode treatment groups: measuring health care with meaningful episodes of care. 2012.

37. Peterson C, Grosse SD, Dunn A. A practical guide to episode groupers for cost-of-illness analysis in health services research. SAGE Open Med. 2019;7:2050312119840200. doi:10.1177/2050312119840200

38. Viera AJ. Odds ratios and risk ratios: what’s the difference and why does it matter? South Med J. Jul 2008;101(7):730–4. doi:10.1097/SMJ.0b013e31817a7ee4

