## Supplement - State Summary for "Neck pain service utilization and costs: association with number of visits of chiropractic manipulation, active care, manual therapy, or acupuncture. A retrospective cohort study"

### Supplement - Episode count by State of individual's home address

| State | Total # | Total % | DC | PT | LAc | State | Total # | Total % | DC | PT | LAc |
|---|---|---|---|---|---|---|---|---|---|---|---|
| Total | 91805 | 100.0% | 87115 | 2617 | 2073 | KY | 989 | 1.1% | 965 | 23 | 1 |
| TX | 10252 | 11.2% | 10022 | 124 | 106 | UT | 908 | 1.0% | 873 | 29 | 6 |
| FL | 7015 | 7.6% | 6767 | 137 | 111 | AR | 798 | 0.9% | 780 | 17 | 1 |
| MO | 5695 | 6.2% | 5663 | 22 | 10 | MA | 730 | 0.8% | 679 | 32 | 19 |
| WI | 5149 | 5.6% | 5013 | 128 | 8 | CT | 721 | 0.8% | 665 | 40 | 16 |
| MN | 4677 | 5.1% | 4412 | 201 | 64 | SC | 588 | 0.6% | 568 | 18 | 2 |
| OH | 4421 | 4.8% | 4318 | 89 | 14 | MS | 544 | 0.6% | 531 | 13 | 0 |
| CA | 4410 | 4.8% | 3722 | 161 | 527 | RI | 527 | 0.6% | 438 | 44 | 45 |
| IL | 4045 | 4.4% | 3851 | 156 | 38 | NV | 445 | 0.5% | 423 | 14 | 8 |
| CO | 3072 | 3.4% | 2824 | 159 | 89 | ND | 351 | 0.4% | 348 | 3 | 0 |
| NC | 2918 | 3.2% | 2797 | 110 | 11 | NM | 320 | 0.4% | 262 | 3 | 55 |
| IA | 2674 | 2.9% | 2644 | 29 | 1 | AL | 280 | 0.3% | 267 | 11 | 2 |
| AZ | 2636 | 2.9% | 2570 | 61 | 5 | DC | 247 | 0.3% | 208 | 15 | 24 |
| GA | 2568 | 2.8% | 2477 | 80 | 11 | SD | 213 | 0.2% | 212 | 1 | 0 |
| VA | 2281 | 2.5% | 2104 | 134 | 43 | NH | 163 | 0.2% | 152 | 7 | 4 |
| NY | 2227 | 2.4% | 1797 | 251 | 179 | WV | 156 | 0.2% | 152 | 4 | 0 |
| WA | 2223 | 2.4% | 1960 | 45 | 218 | ID | 134 | 0.2% | 130 | 4 | 0 |
| NE | 2197 | 2.4% | 2176 | 18 | 3 | ME | 131 | 0.1% | 125 | 4 | 2 |
| PA | 1946 | 2.1% | 1900 | 32 | 14 | VI | 109 | 0.1% | 106 | 3 | 0 |
| IN | 1744 | 1.9% | 1690 | 54 | 0 | WY | 86 | 0.1% | 79 | 6 | 1 |
| MD | 1710 | 1.9% | 1471 | 113 | 126 | DE | 82 | 0.1% | 79 | 2 | 1 |
| TN | 1485 | 1.6% | 1461 | 22 | 2 | MT | 77 | 0.1% | 72 | 4 | 1 |
| OR | 1360 | 1.5% | 1099 | 35 | 226 | VT | 19 | 0.0% | 18 | 0 | 1 |
| NJ | 1244 | 1.4% | 1138 | 50 | 56 | HI | 15 | 0.0% | 12 | 3 | 0 |
| KS | 1238 | 1.4% | 1225 | 11 | 2 | AK | 14 | 0.0% | 13 | 1 | 0 |
| OK | 1163 | 1.3% | 1150 | 12 | 1 | PR | 7 | 0.0% | 7 | 0 | 0 |
| LA | 1162 | 1.3% | 1134 | 26 | 2 | Unknown | 619 | 0.7% | 581 | 24 | 14 |
| MI | 1020 | 1.1% | 985 | 32 | 3 | | | | | | |
