## Supplement 1 - Distribution of Episodes for "Neck pain service utilization and costs: association with number of visits of chiropractic manipulation, active care, manual therapy, or acupuncture. A retrospective cohort study"

| Supplement 1 - Single episode non-surgical neck pain episode distribution by type of initial contact health care provider (HCP) and number of visits of service | | | | | | | | | |
|---|---|---|---|---|---|---|---|---|---|
| HCP Type | Service Type | # of Visits of Service | | | | | | | |
| | | Service Not Provided | 1 to 3 visits (reference) | 4 to 6 visits | 7 to 9 visits | 10 to 12 visits | 13 to 15 visits | 16+ visits | Total |
| Episode Count | | | | | | | | | |
| DC | CMT | 6010 | 40373 | 14447 | 7348 | 5460 | 3357 | 7455 | 84450 |
| | AC | 58481 | 14804 | 4556 | 2256 | 1453 | 864 | 2036 | 84450 |
| | MT | 68115 | 10159 | 2781 | 1263 | 857 | 454 | 821 | 84450 |
| | Acu | 83792 | 371 | 117 | 51 | 40 | 15 | 64 | 84450 |
| PT | AC | 128 | 821 | 488 | 363 | 234 | 135 | 410 | 2579 |
| | MT | 481 | 903 | 502 | 323 | 149 | 90 | 131 | 2579 |
| | CMT | 2302 | 106 | 58 | 31 | 25 | 13 | 44 | 2579 |
| | Acu | 2529 | 14 | 14 | 5 | 7 | 2 | 8 | 2579 |
| LAc | Acu | 177 | 555 | 559 | 193 | 210 | 82 | 254 | 2030 |
| | MT | 1139 | 539 | 184 | 70 | 36 | 19 | 43 | 2030 |
| | AC | 1688 | 173 | 76 | 29 | 19 | 11 | 34 | 2030 |
| | CMT | 1796 | 102 | 48 | 24 | 16 | 17 | 27 | 2030 |
| % of Episodes | | | | | | | | | |
| DC | **CMT** | **7.1%** | 47.8% | 17.1% | 8.7% | 6.5% | 4.0% | 8.8% | 100.0% |
| | AC | 69.2% | 17.5% | 5.4% | 2.7% | 1.7% | 1.0% | 2.4% | 100.0% |
| | MT | 80.7% | 12.0% | 3.3% | 1.5% | 1.0% | 0.5% | 1.0% | 100.0% |
| | Acu | 99.2% | 0.4% | 0.1% | 0.1% | 0.0% | 0.0% | 0.1% | 100.0% |
| PT | **AC** | **5.0%** | 31.8% | 18.9% | 14.1% | 9.1% | 5.2% | 15.9% | 100.0% |
| | **MT** | **18.7%** | 35.0% | 19.5% | 12.5% | 5.8% | 3.5% | 5.1% | 100.0% |
| | CMT | 89.3% | 4.1% | 2.2% | 1.2% | 1.0% | 0.5% | 1.7% | 100.0% |
| | Acu | 98.1% | 0.5% | 0.5% | 0.2% | 0.3% | 0.1% | 0.3% | 100.0% |
| LAc | **Acu** | **8.7%** | 27.3% | 27.5% | 9.5% | 10.3% | 4.0% | 12.5% | 100.0% |
| | MT | 56.1% | 26.6% | 9.1% | 3.4% | 1.8% | 0.9% | 2.1% | 100.0% |
| | AC | 83.2% | 8.5% | 3.7% | 1.4% | 0.9% | 0.5% | 1.7% | 100.0% |
| | CMT | 88.5% | 5.0% | 2.4% | 1.2% | 0.8% | 0.8% | 1.3% | 100.0% |

DC=Doctor of Chiropractic, PT=Physical Therapist, LAc=Licensed Acupuncturist, CMT=Chiropractic Manipulative Treatment, AC=Active Care, MT=Manual Therapy, Acu=Acupuncture
