## Supplement 2 - Cohort Summary for "Neck pain service utilization and costs: association with number of visits of chiropractic manipulation, active care, manual therapy, or acupuncture. A retrospective cohort study"

| Supplement 2 - Non-surgical single episode neck pain cohort characteristics for individuals initially contacting a Chiropractor (DC), Physical Therapist (PT), or Licensed Acupuncturists (LAc) | | | |
|---|---|---|---|
| | DC | PT | LAc |
| # Unique Health Care Providers (HCP) | 19387 | 1828 | 1153 |
| Episodes | 87115 | 2617 | 2073 |
| Individuals | 87115 | 2617 | 2073 |
| Total Cost | 34911677 | 2874276 | 1364991 |
| Individuals - % or (Median (Q1,Q3)) | | | |
| % Female | 58.8% | 65.1% | 70.5% |
| Age | 41 (31, 51) | 47 (36, 56) | 40 (33, 49) |
| ERG® Risk Score | 1.0 (0.5, 2.1) | 1.9 (0.9, 3.5) | 1.1 (0.5, 2.3) |
| Individual Home Address 5 Digit Zip Code Population Attributes - (Median (Q1,Q3)) | | | |
| % Non-Hispanic White (NHW) | 76.8% (60.1%, 88.1%) | 73.1% (54.1%, 84.4%) | 63.8% (43.0%, 78.0%) |
| Area Deprivation Index (ADI) | 43 (28, 60) | 33 (19, 50) | 23 (13, 36) |
| Household Adjusted Gross Income (AGI) | 67,793 (53,596, 93,292) | 77,563 (57,639, 113,129) | 91,222 (64,727, 134,555) |
| DC per 1000 Population | 0.28 (0.13, 0.50) | 0.26 (0.12, 0.48) | 0.27 (0.13, 0.51) |
| PT per 1000 Population | 0.19 (0.05, 0.45) | 0.26 (0.09, 0.55) | 0.26 (0.09, 0.59) |
| LAc per 1000 Population | 0.00 (0.00, 0.03) | 0.00 (0.00, 0.06) | 0.06 (0.00, 0.19) |
| Episode Attributes - % or (Median (Q1,Q3)(Minimum)) | | | |
| % Without CMT, AC, MT and Acu | 3.1% | 1.5% | 2.1% |
| Episode Cost | $180 (70, 450) | $604 (308, 1292) | $330 (153, 698) |
| # of Different HCP Seen | 1 (1, 2) | 2 (1, 4) | 1 (1, 2) |
| Episode Duration - Days | 27 (3, 101) (1) | 45 (22, 106) (1) | 29 (5, 66) (1) |
| Clean Period Before Episode - Days | 668 (458, 874) (91) | 670 (457, 865) (91) | 698 (476, 900) (93) |
| Clean Period After Episode - Days | 405 (239, 638) (61) | 399 (237, 648) (61) | 405 (237, 637) (61) |

CMT=Chiropractic Manipulative Treatment, AC=Active Care, MT=Manual Therapy, Acu=Acupuncture
