## Supplement 3 - Cohort Details for "Neck pain service utilization and costs: association with number of visits of chiropractic manipulation, active care, manual therapy, or acupuncture. A retrospective cohort study"

**Supplement 3 - Single episode non-surgical neck pain cohort characteristics by type of initial contact health care provider and number of visits of select first line service**

| % or Median (Q1, Q3) | Service Not Provided | 1 to 3 visits (reference) | 4 to 6 visits | 7 to 9 visits | 10 to 12 visits | 13 to 15 visits | 16+ visits | Total |
|---|---|---|---|---|---|---|---|---|
| **Initial Contact With Chiropractor (DC) - # of Visits of Chiropractic Manipulative Treatment (CMT)** | | | | | | | | |
| # of DCs | 594 | 14011 | 8448 | 5234 | 4043 | 2744 | 4802 | 19195 |
| % of DCs | 3.1% | 73.0% | 44.0% | 27.3% | 21.1% | 14.3% | 25.0% | 100.0% |
| Episodes/Individuals | 6010 | 40373 | 14447 | 7348 | 5460 | 3357 | 7455 | 84450 |
| % of Episodes/Individuals | 7.1% | 47.8% | 17.1% | 8.7% | 6.5% | 4.0% | 8.8% | 100.0% |
| Individuals - % Female | 59.8% | 56.2% | 58.8% | 60.6% | 61.7% | 62.9% | 65.3% | 58.7% |
| Individuals - Age | 40 (31, 49) | 39 (30, 50) | 42 (32, 52) | 42 (32, 52) | 42 (33, 53) | 43 (33, 53) | 43 (34, 53) | 41 (31, 51) |
| Individuals - ERG® Risk Score | 1.1 (0.5, 2.2) | 0.9 (0.4, 2.0) | 1.0 (0.5, 2.1) | 1.1 (0.6, 2.3) | 1.1 (0.6, 2.2) | 1.2 (0.6, 2.3) | 1.2 (0.6, 2.4) | 1.0 (0.5, 2.1) |
| Zip Code - % non-Hispanic White | 68% (50, 82) | 78% (61, 89) | 78% (61, 88) | 78% (61, 88) | 77% (61, 88) | 77% (61, 88) | 77% (61, 88) | 77% (60, 88) |
| Zip Code - Area Deprivation Index | 43 (27, 59) | 44 (29, 60) | 42 (26, 59) | 43 (27, 59) | 42 (27, 58) | 42 (27, 58) | 41 (26, 57) | 43 (28, 60) |
| Zip Code - DCs per 1000 | 0.23 (0.10, 0.43) | 0.28 (0.12, 0.50) | 0.28 (0.13, 0.51) | 0.28 (0.13, 0.51) | 0.28 (0.13, 0.51) | 0.27 (0.13, 0.49) | 0.29 (0.14, 0.52) | 0.28 (0.13, 0.50) |
| Zip Code - PTs per 1000 | 0.16 (0.04, 0.38) | 0.19 (0.05, 0.45) | 0.20 (0.06, 0.46) | 0.20 (0.06, 0.47) | 0.21 (0.06, 0.47) | 0.20 (0.05, 0.47) | 0.20 (0.06, 0.47) | 0.19 (0.05, 0.45) |
| Zip Code - LAcs per 1000 | 0.00 (0.00, 0.03) | 0.00 (0.00, 0.02) | 0.00 (0.00, 0.03) | 0.00 (0.00, 0.03) | 0.00 (0.00, 0.03) | 0.00 (0.00, 0.03) | 0.00 (0.00, 0.03) | 0.00 (0.00, 0.03) |
| Clean Period Before Episode - days | 718 (485, 903) | 717 (473, 912) | 670 (461, 871) | 641 (443, 844) | 587 (409, 821) | 563 (394, 791) | 521 (347, 733) | 667 (457, 873) |
| Clean Period After Episode - days | 419 (244, 657) | 431 (244, 679) | 414 (237, 639) | 392 (230, 595) | 388 (244, 578) | 378 (232, 536) | 375 (236, 508) | 405 (241, 638) |
| **Initial Contact With Physical Therapist (PT) - # of Visits of Active Care (AC)** | | | | | | | | |
| # of PTs | 75 | 688 | 418 | 330 | 208 | 120 | 322 | 1800 |
| % of PTs | 4.2% | 38.2% | 23.2% | 18.3% | 11.6% | 6.7% | 17.9% | 100.0% |
| Episodes/Individuals | 128 | 821 | 488 | 363 | 234 | 135 | 410 | 2579 |
| % of Episodes/Individuals | 5.0% | 31.8% | 18.9% | 14.1% | 9.1% | 5.2% | 15.9% | 100.0% |
| Individuals - % Female | 67.2% | 63.0% | 66.0% | 69.7% | 68.4% | 66.7% | 60.5% | 65.0% |
| Individuals - Age | 46 (36, 57) | 46 (35, 55) | 47 (36, 55) | 49 (36, 56) | 48 (36, 56) | 45 (34, 56) | 47 (37, 55) | 47 (36, 56) |
| Individuals - ERG® Risk Score | 2.3 (1.0, 3.9) | 1.7 (0.9, 3.5) | 1.9 (0.9, 3.4) | 1.8 (1.0, 3.3) | 1.9 (1.0, 3.5) | 1.7 (0.9, 3.3) | 1.9 (1.1, 3.8) | 1.8 (0.9, 3.5) |
| Zip Code - % non-Hispanic White | 74% (52, 85) | 74% (59, 85) | 73% (53, 84) | 71% (50, 84) | 73% (54, 83) | 70% (50, 83) | 73% (56, 85) | 73% (54, 84) |
| Zip Code - Area Deprivation Index | 32 (19, 56) | 34 (20, 52) | 35 (22, 50) | 34 (20, 50) | 32 (18, 49) | 35 (20, 54) | 28 (15, 45) | 33 (19, 50) |
| Zip Code - DCs per 1000 | 0.25 (0.12, 0.46) | 0.26 (0.12, 0.48) | 0.27 (0.11, 0.48) | 0.23 (0.11, 0.46) | 0.25 (0.12, 0.47) | 0.24 (0.12, 0.50) | 0.27 (0.12, 0.49) | 0.26 (0.12, 0.48) |
| Zip Code - PTs per 1000 | 0.27 (0.10, 0.56) | 0.26 (0.09, 0.59) | 0.25 (0.08, 0.54) | 0.26 (0.08, 0.55) | 0.24 (0.08, 0.52) | 0.23 (0.07, 0.46) | 0.31 (0.12, 0.60) | 0.26 (0.09, 0.56) |
| Zip Code - LAcs per 1000 | 0.00 (0.00, 0.04) | 0.00 (0.00, 0.05) | 0.00 (0.00, 0.06) | 0.00 (0.00, 0.05) | 0.00 (0.00, 0.05) | 0.00 (0.00, 0.06) | 0.00 (0.00, 0.07) | 0.00 (0.00, 0.06) |
| Clean Period Before Episode - days | 678 (452, 867) | 717 (468, 892) | 684 (457, 852) | 675 (458, 850) | 646 (457, 864) | 583 (384, 827) | 613 (348, 824) | 670 (457, 865) |
| Clean Period After Episode - days | 388 (192, 623) | 395 (238, 655) | 406 (243, 664) | 420 (242, 658) | 394 (233, 642) | 425 (271, 677) | 389 (236, 593) | 398 (237, 648) |
| **Initial Contact With Physical Therapist (PT) - # of Visits of Manual Therapy (MT)** | | | | | | | | |
| # of PTs | 305 | 735 | 421 | 286 | 135 | 84 | 122 | 1800 |
| % of PTs | 16.9% | 40.8% | 23.4% | 15.9% | 7.5% | 4.7% | 6.8% | 100.0% |
| Episodes/Individuals | 481 | 903 | 502 | 323 | 149 | 90 | 131 | 2579 |
| % of Episodes/Individuals | 18.7% | 35.0% | 19.5% | 12.5% | 5.8% | 3.5% | 5.1% | 100.0% |
| Individuals - % Female | 62.4% | 63.2% | 69.3% | 65.9% | 62.4% | 65.6% | 70.2% | 65.0% |
| Individuals - Age | 46 (35, 55) | 47 (36, 56) | 47 (35, 55) | 49 (37, 55) | 48 (41, 58) | 47 (36, 57) | 47 (35, 54) | 47 (36, 56) |
| Individuals - ERG® Risk Score | 1.8 (1.0, 3.8) | 1.8 (0.9, 3.4) | 1.9 (1.0, 3.4) | 1.7 (0.7, 3.2) | 1.8 (0.9, 4.1) | 1.9 (1.0, 4.2) | 2.5 (1.4, 4.0) | 1.8 (0.9, 3.5) |
| Zip Code - % non-Hispanic White | 72% (53, 84) | 74% (58, 86) | 71% (50, 82) | 75% (53, 85) | 72% (57, 85) | 70% (45, 80) | 77% (61, 86) | 73% (54, 84) |
| Zip Code - Area Deprivation Index | 37 (22, 55) | 34 (19, 49) | 32 (20, 49) | 32 (18, 46) | 35 (16, 50) | 30 (13, 49) | 28 (15, 45) | 33 (19, 50) |
| Zip Code - DCs per 1000 | 0.27 (0.13, 0.49) | 0.26 (0.12, 0.46) | 0.24 (0.11, 0.49) | 0.27 (0.13, 0.50) | 0.26 (0.12, 0.43) | 0.20 (0.10, 0.44) | 0.28 (0.13, 0.53) | 0.26 (0.12, 0.48) |
| Zip Code - PTs per 1000 | 0.26 (0.08, 0.54) | 0.24 (0.08, 0.55) | 0.27 (0.09, 0.55) | 0.27 (0.09, 0.58) | 0.24 (0.08, 0.53) | 0.29 (0.07, 0.50) | 0.35 (0.12, 0.72) | 0.26 (0.09, 0.56) |
| Zip Code - LAcs per 1000 | 0.00 (0.00, 0.05) | 0.00 (0.00, 0.05) | 0.00 (0.00, 0.06) | 0.00 (0.00, 0.05) | 0.00 (0.00, 0.06) | 0.03 (0.00, 0.07) | 0.00 (0.00, 0.09) | 0.00 (0.00, 0.06) |
| Clean Period Before Episode - days | 674 (454, 873) | 710 (465, 892) | 680 (458, 847) | 649 (441, 850) | 636 (457, 825) | 581 (260, 825) | 495 (228, 784) | 670 (457, 865) |
| Clean Period After Episode - days | 402 (263, 663) | 395 (225, 643) | 406 (231, 652) | 432 (250, 678) | 376 (242, 579) | 444 (245, 606) | 379 (247, 590) | 398 (237, 648) |
| **Initial Contact with Licensed Acupuncturist (LAc) - # of Visits of Acupuncture (Acu)** | | | | | | | | |
| # of LAcs | 57 | 437 | 443 | 173 | 184 | 76 | 208 | 1142 |
| % of LAcs | 5.0% | 38.3% | 38.8% | 15.1% | 16.1% | 6.7% | 18.2% | 100.0% |
| Episodes/Individuals | 177 | 555 | 559 | 193 | 210 | 82 | 254 | 2030 |
| % of Episodes/Individuals | 8.7% | 27.3% | 27.5% | 9.5% | 10.3% | 4.0% | 12.5% | 100.0% |
| Individuals - % Female | 74.0% | 69.2% | 70.7% | 67.4% | 74.8% | 74.4% | 68.9% | 70.6% |
| Individuals - Age | 41 (35, 50) | 40 (32, 49) | 39 (32, 48) | 41 (34, 50) | 41 (33, 50) | 39 (32, 49) | 42 (34, 51) | 40 (33, 49) |
| Individuals - ERG® Risk Score | 1.3 (0.7, 2.8) | 1.1 (0.4, 2.4) | 0.9 (0.4, 1.9) | 1.0 (0.5, 2.1) | 1.2 (0.5, 2.4) | 1.1 (0.3, 2.4) | 1.4 (0.6, 2.6) | 1.1 (0.5, 2.3) |
| Zip Code - % non-Hispanic White | 61% (37, 74) | 64% (44, 78) | 66% (45, 79) | 66% (48, 78) | 62 (41, 78) | 61% (38, 76) | 62% (41, 76) | 64% (43, 78) |
| Zip Code - Area Deprivation Index | 27 (17, 46) | 24 (13, 36) | 24 (13, 37) | 22 (11, 35) | 21 (13, 34) | 21 (13, 35) | 20 (11, 32) | 23 (13, 36) |
| Zip Code - DCs per 1000 | 0.24 (0.10, 0.48) | 0.29 (0.13, 0.51) | 0.29 (0.13, 0.51) | 0.29 (0.13, 0.50) | 0.26 (0.12, 0.49) | 0.25 (0.11, 0.53) | 0.26 (0.12, 0.51) | 0.27 (0.13, 0.51) |
| Zip Code - PTs per 1000 | 0.20 (0.06, 0.48) | 0.26 (0.10, 0.65) | 0.26 (0.09, 0.61) | 0.25 (0.08, 0.57) | 0.27 (0.10, 0.57) | 0.32 (0.10, 0.60) | 0.25 (0.11, 0.58) | 0.26 (0.09, 0.59) |
| Zip Code - LAcs per 1000 | 0.04 (0.00, 0.16) | 0.07 (0.00, 0.23) | 0.06 (0.00, 0.19) | 0.06 (0.00, 0.20) | 0.07 (0.02, 0.18) | 0.07 (0.01, 0.23) | 0.06 (0.00, 0.17) | 0.06 (0.00, 0.19) |
| Clean Period Before Episode - days | 661 (440, 866) | 719 (482, 908) | 730 (514, 920) | 739 (493, 914) | 648 (450, 864) | 708 (485, 910) | 618 (412, 821) | 699 (479, 900) |
| Clean Period After Episode - days | 418 (257, 645) | 426 (258, 662) | 423 (230, 624) | 389 (225, 635) | 412 (219, 634) | 362 (191, 628) | 374 (225, 553) | 403 (237, 634) |

Cells with red text denote that the effect of provider type on service usage was found not to be significantly different from that of PCP-reference (Mann-Whitney U $p > 0.001$)

Cells with black text denote that the effect of provider type on service usage was found to be significantly different from that of PCP-reference (Mann-Whitney U $p < 0.001$)
